# Wastewater testing during the South African 2022-2023 measles outbreak demonstrates the potential of environmental surveillance to support measles elimination

**DOI:** 10.1101/2024.09.01.24312904

**Authors:** Nkosenhle Ndlovu, Kerrigan M. McCarthy, Victor Vusi Mabasa, Chenoa Sankar, Nosihle Msomi, Emmanuel Phalane, Natasha Singh, Sipho Gwala, Fiona Els, Mokgaetji Macheke, Sibonginkosi Maposa, Mukhlid Yousif

## Abstract

**Background:** Surveillance is a key component of the WHO 2030 measles elimination strategy. Wastewater and environmental surveillance (WES) has successfully supported polio surveillance, however, it has not been applied to measles control.

**Aim:** We developed a measles virus (MeV) digital-PCR (dPCR) assay using WHO recommended clinical primer-probes and applied it to retained wastewater samples from national SARS-CoV-2 wastewater sentinel surveillance sites. We compared results with national clinical measles surveillance data.

**Setting:** Wastewater concentrates were collected from 47 SARS-CoV-2 wastewater surveillance sites across South Africa.

**Methods:** We determined the limit of detection using the assay on serial dilutions of MeV positive controls. Subsequently we conducted batch testing on wastewater concentrates retained at -20°C for up to 15 months and compared wastewater results with national laboratory-confirmed measles case data by district and epidemiological week.

**Results:** Our assay identified 43/2149 (2%) samples containing MeV RNA in concentrations ranging from 1.97 – 165.8 genome copies/mL. Amongst 27 week-district instances where at least one MeV positive wastewater sample was detected, no clinical cases were detected in 13 (48%) of these.

**Conclusion:** Despite likely RNA degradation, it was possible to detect MeV in wastewater samples in districts where clinical surveillance failed to identify cases.

**Contribution:** WES has evident potential to strengthen surveillance in support of the measles elimination agenda. With immediate processing and improved wastewater RNA concentration methods, WES sensitivity will likely increase.

**Funding:** This work was funded by BMGF (INV-049271)

## Introduction

Measles, a highly infectious, vaccine-preventable viral infection, is targeted by the World Health Organization (WHO) for elimination by 2030[1]. Measles incidence and mortality have declined dramatically since the year 2000 due to the increase in global vaccination coverage from 72% in 2000 to 83% in 2022, despite a transient decline in 2021 due to the COVID-19 pandemic. Global measles indicators have improved with measles deaths falling between 2000 and 2022 from 1,072,800 to 136,200 deaths and measles incidence dropping from 145 cases per million, to 29 cases per million [2]. Despite these improvements, measles is still a leading cause of death in children under the age of 5 years in low- and middle-income countries[2]. In support of the 2021-2030 strategic framework for measles and rubella elimination, ongoing global efforts are in place to control viral transmission[1]. Efforts are targeted at ensuring high vaccination coverage and implementation of sensitive surveillance systems[1]. Interventions include the provision of two doses of measles vaccine before the age of 5 years as part of the expanded programme of immunisation (EPI), supplementary immunisation activities (SIAs) every 4-5 years where routine vaccination coverage does not achieve 95% coverage, fever-rash surveillance, and rapid, outbreak response[1]. In South Africa, intermittent measles outbreaks have continued to occur over the last 25 years, the largest of which led to over 21,000 laboratory-confirmed cases 2009-2011[3]. Most recently, an outbreak of over 1,383 cases commenced in 2022 following apparent importation of the B3 genotype from neighbouring countries.

As measles elimination targets and dates approach, the WHO and national public health authorities rely increasingly on measles surveillance to support program monitoring[1]. Theoretically, measles surveillance should be highly sensitive, as asymptomatic infection does not occur, and a robust, though non-specific case definition is easily applied[4]. However, four factors may limit the sensitivity and/or utility of clinical measles surveillance. Firstly, health system factors including clinician awareness, propensity to test and notify, financial allocations for testing, and submission of diagnostic specimens often limit submission of diagnostic specimens[5]. Secondly, in our experience, patients and their caregivers may not seek medical care if their clinical presentation is uncomplicated as may occur in the context of older children, and recognized outbreaks. Thirdly, incomplete provision of clinical data (such as date of rash onset and vaccination history) leads to challenges in case classification, contact tracing and outbreak response[6]. Fourthly, urine or throat swabs are less frequently submitted for polymerase chain reaction (PCR) testing, rendering PCR detection and genomic sequencing, and thus case classification especially difficult during outbreak contexts[7].

Wastewater and environmental surveillance (WES) has been a component of the Global Polio Elimination Initiative since 2003. In recent years, WES has been applied across the globe to provide complementary surveillance data during the SARS-CoV-2 pandemic, the multicountry mpox clade II outbreak[8], and other pathogens. Regarding measles surveillance, the earliest application of WES was conducted by Benschkop *et al* who detected MeV in 6/56 (10.7%) wastewater samples collected for polio surveillance during a measles outbreak in a vaccine-hesitant community 2013[9]. A number of laboratory protocols for measles detection have been developed[10–14] and applied to outbreak contexts retrospectively[15] and prospectively[12,16]. However, to date measles WES is yet to be implemented systematically as part of routine programmatic surveillance.

In this paper, we describe the development, optimisation and validation of a reverse transcriptase digital PCR (RT-dPCR) assay for detection and quantification of MeV in wastewater, which we applied retrospectively to wastewater samples from a national sentinel surveillance network. As wastewater samples were collected during the ongoing national measles outbreak that began in October 2022, we compared wastewater findings with clinical surveillance data to explore the role of WES as a complementary surveillance tool for programmatic purposes.

## Methods

### Study design and setting

In 2020, the National Institute of Communicable Diseases (NICD) expanded polio testing at 18 WES sentinel sites to include SARS-CoV-2 across nine provinces of South Africa[17]. Over time, and dependent on funding, the number and location of sites changed. Presently, the NICD coordinates a national WES sentinel surveillance network comprising sampling sites at 28 wastewater treatment plants (WWTPs) across nine South African provinces and border transit points, and a network of 19 community sampling sites located within the catchment areas of three large WWTPs in the densely populated Gauteng Province (Figure 1).

**Figure 1.**
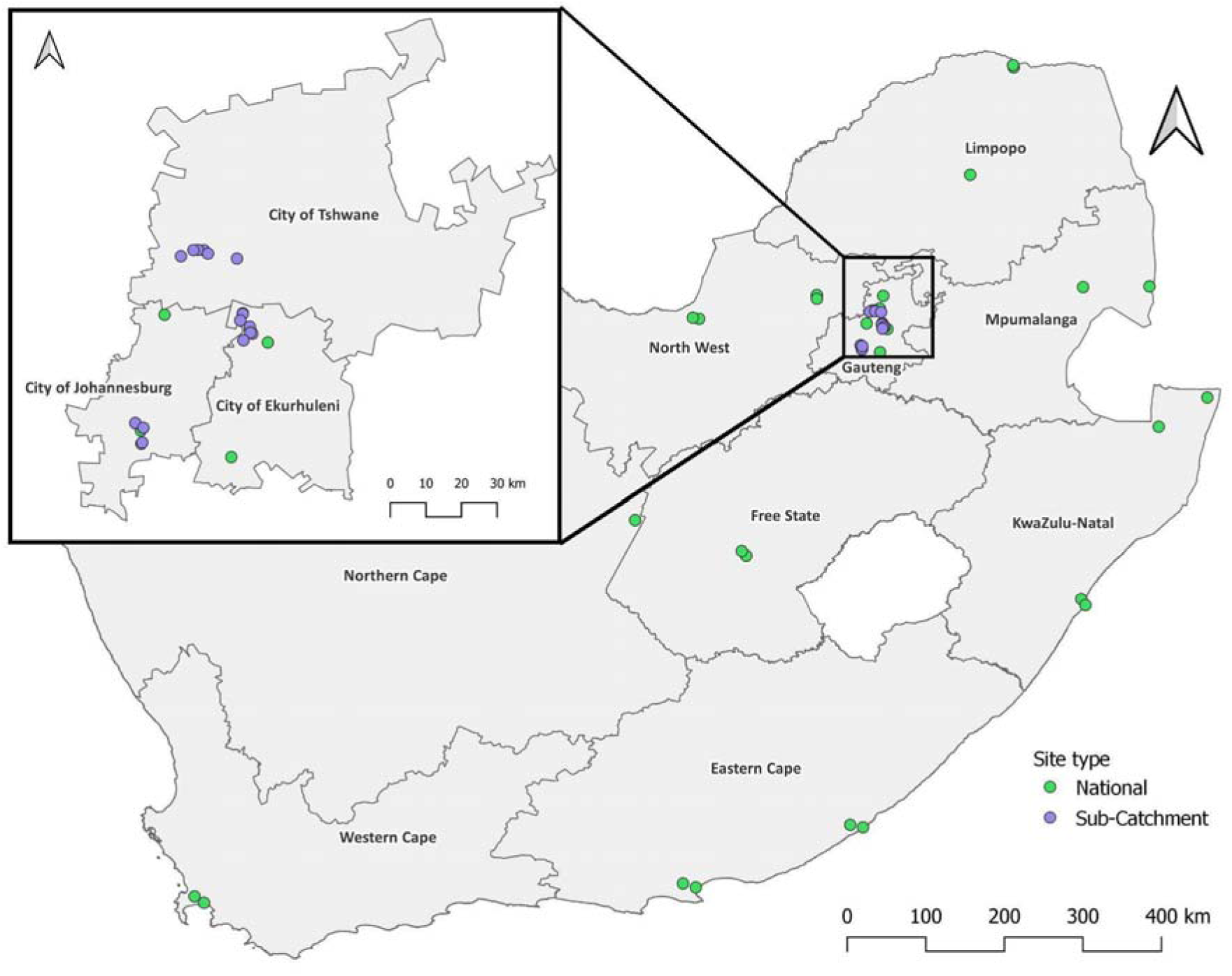
Sampling locations at South African wastewater treatment plants (national sentinel surveillance sites) and in-line sewer sampling sites (sub-catchment areas in City of Tshwane, Johannesburg and Ekurhuleni, inset). Figure created using QGIS (version 3.42.3) with opensource shapefiles from https://www.demarcation.org.za/

### Laboratory methods

#### Wastewater sample collection, concentration and storage

Grab samples of one litre were collected monthly, weekly or biweekly from collection sites by immersion of a Virocide® treated bucket lowered into the middle of the sewage stream, at approximately the same time of day. Samples were decanted into one litre plastic screwtop bottles and transported on ice to the NICD within 24 hours of collection. On receipt, samples were refrigerated at 4°C, and within 24-48 hours, 200mL of raw sewage was centrifuged at 4650 x g at 4°C for 30 minutes to clarify the sample. Then 70mL of supernatant was centrifuged at 3500 x g for 15 minutes through a Centricon^®^ Plus-70 centrifugal ultra-filter (Merck Millipore, Tullagreen, Ireland). Material retained in the filter was eluted to a concentrate volume of approximately 1 mL. Concentrates were processed immediately for SARS-CoV-2, and the balance was retained at -20 °C for extraction and PCR once MeV and other assays were developed and validated.

#### Primers and probes for measles and pepper mild mottle virus (PMMoV) detection

Primers and probes specific for MeV and pepper mild mottle virus (PMMoV) were identified from previously described methods[18,19] (Table 1). We targeted a fragment of the N gene and designed ‘MeV-WT’ and genotype A-specific primers and a probe to differentiate wild-type MeV genotypes dominant in recent South African outbreaks (B3, D8 and H1) from vaccine strain (genotype A) (Table 1, Figure 2).

**Figure 2.**
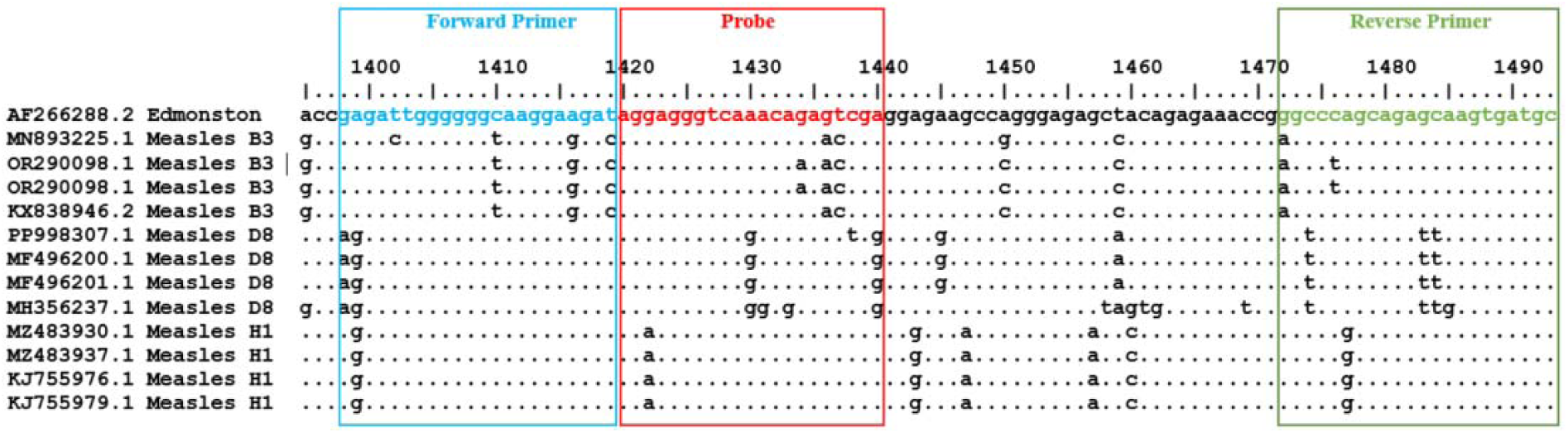
Primers and probe design to differentiate measles virus vaccine strain (Edmonston) from wild genotypes (B3, D8, and H1) using the N gene.

**Table 1.**
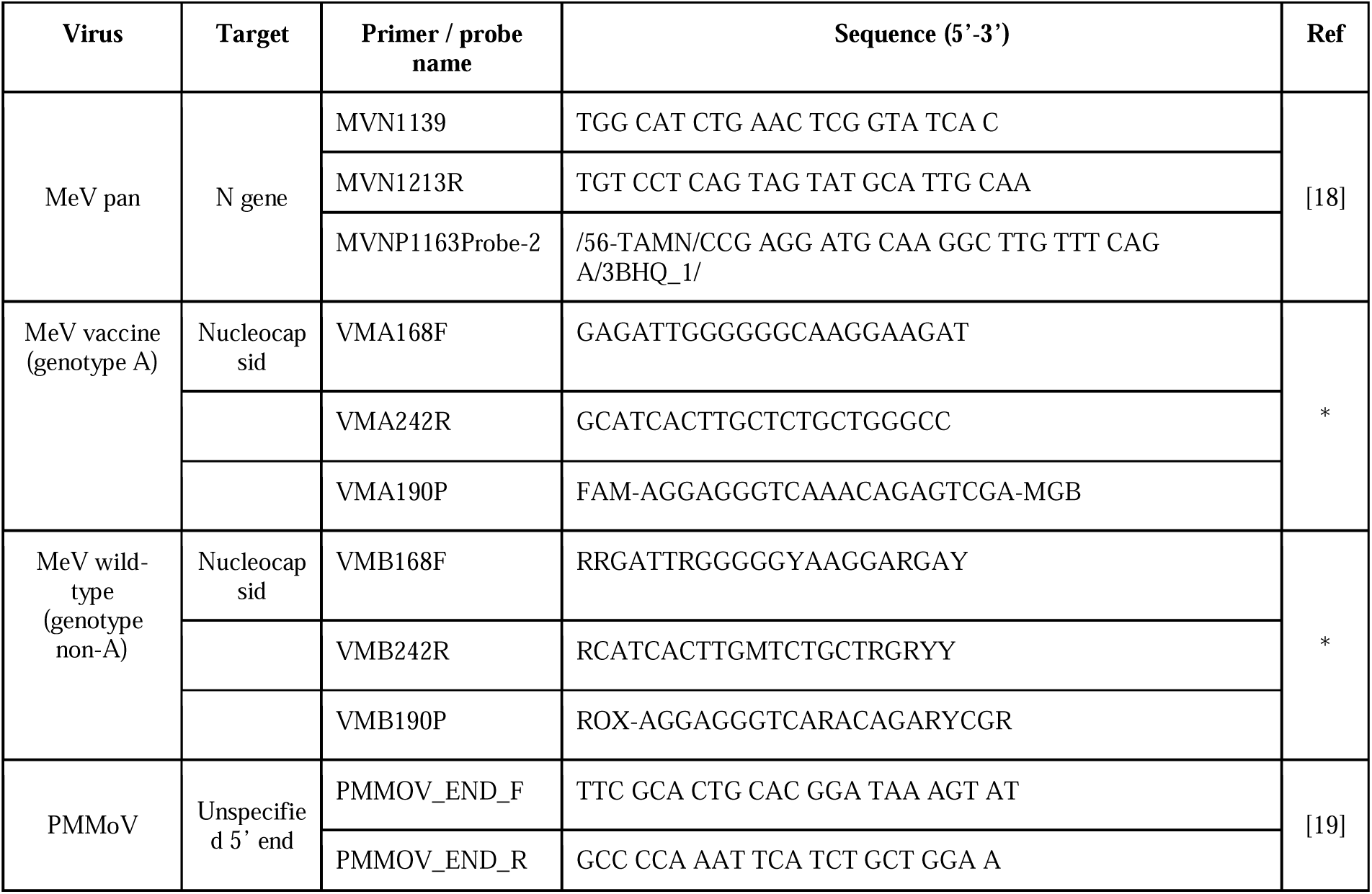
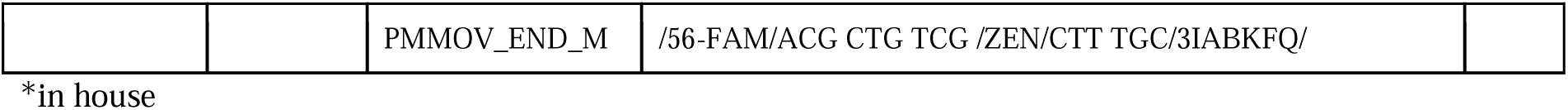
Primers, probes and source references used for PCR detection of measles virus (MeV) and pepper mild mottle virus (PMMoV) in wastewater samples.

Using the measles RNA-positive control provided by the Global Measles Reference Laboratory Network (GMRLN), the commercially available OMZYTA® MMR live attenuated vaccine (MSD (Pty) Ltd), derived from Enders’ attenuated Edmonston strain, and RNA from clinical samples collected during an outbreak in South Africa in 2022 as positive controls, we performed dPCR assays in replicates of five using the method described below using the MeV pan, MeV-WT and genotype A-specific primer-probe combinations to determine relative specificity.

#### Digital PCR optimisation and determination of limit of detection

We determined the limit of detection (LoD) (defined as the concentration in genome copies/uL of the lowest dilution that is theoretically detectable when using optimized methods) with a single- and duplex RT-dPCR assay using pan-MeV primers and probe with and without previously described rubella virus (RuV) primers and probe[20]. In triplicate, we serially diluted the MeV RNA-positive control provided by the GMRLN in 14 two-fold dilutions (neat to 1:16,384) and subjected these to the QIAcuity OneStep Advanced Probe RT-dPCR assay (Qiagen, Hilden, Germany) on QIAcuity® One, 5plex System dPCR platform (Qiagen) using 96-well 8.5k nanoplates. The pan-MeV RT-dPCR master mix contained 3 µL of 4× QIAcuity One-Step Advanced Probe Master Mix, 0.12 µL of 100× One-Step Advanced RT-Mix, 0.6 µL of 20× primer-probe mix MeV (TAMRA), 0.6 µL of 20× RuV primer-probe mix (Cy5) and 1 µL of QIAcuity Enhancer GC. The same master mix set-up was used for vaccine-specific primers and probe reactions. Cycling conditions for RT-dPCR were reverse transcription at 50°C for 40 minutes, RT enzyme inactivation at 95°C for 1 minute and 45 cycles of annealing & extension at 60°C.

Each RNA dilution of the positive control was also tested using RT-qPCR conducted on the Applied Biosystems® 7500 Real-Time PCR System (Applied Biosystems, Foster City, CA, USA) using the QIAcuity One-Step Advanced Probe Master Mix. The RT-qPCR master mix contained 4x One-Step Advanced Probe Master Mix 6.25 µL, 100x One-Step Advanced RT-Mix 0.25 µL, 20x primer-probe mix MV (TAMRA) 1.25 µL, 20x primer-probe mix RV (Cy5) 1.25 µL, Enhancer GC 2 µL, RNAse-free water 6 µL.

We defined the assay LoD as one positive partition on the QIAcuity® One, 5plex System dPCR platform (Qiagen), provided that ≥70% of the total partitions are valid. However, when expressed in copies/µL or copies/mL, the LoD represents a range rather than a fixed value, as the calculation is based on a Poisson distribution that incorporates both the number of valid partitions and the number of positive partitions to estimate concentration. We did not determine the LoD in spiked wastewater, however, we are satisfied that a single positive partition, in a raw wastewater sample even in the absence of replicability, is a valid result. Our justification for this is as follows: 1) we observed single positive partitions in serial dilutions of positive control when applying qPCR and dPCR to the same dilutions; 2) replicability in the presence of extremely low concentrations of nucleic acid such as is found in wastewater, is statistically highly unlikely; 3) in subsequent work (manuscript in progress), we have successfully sequenced MeV from wastewater even when a single partition tested positive.

#### Testing of retained wastewater concentrates

Following thawing, over 2,100 retained concentrates from February 2021 (week 7) to March 2024 (week 10) were tested in batches of 92 samples for MeV and PMMoV. Total nucleic acids were extracted from 200 µL of viral concentrate on the 96 KingFisher Flex Purification System (Thermo Fisher Scientific, Waltham, MA USA) using the MagMAX™ Wastewater Ultra Nucleic Acid Isolation kit (Thermo Fisher Scientific) according to the manufacturer’s instructions. Briefly, nucleic acids were captured in 520 µL of lysis binding solution before they were washed in 1ml of Wash 1 and 3 solutions and eluted in 60 µL of elution buffer. We added 8μl of nucleic acids in the dPCR reaction. We used the formula below to determine the genome copy number per milliliter of the raw wastewater sample:

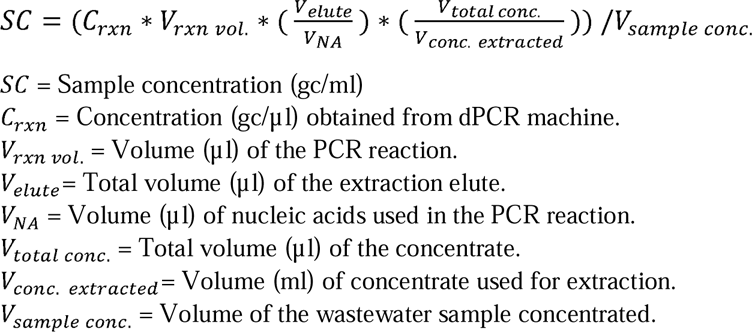

#### Detection of wild-type and genotype A (vaccine strain) in MeV-positive wastewater samples

After identification of MeV positive wastewater samples, we returned to residual retained concentrates from these samples and subjected them to a second extraction and PCR using the QIAcuity® One, 5plex System (Qiagen, Hilden, Germany) as described above, using the in-house vaccine- and wild-type specific primers to identify samples where wild-type and genotype A (vaccine) was present (see Supplement). We used the commercially available OMZYTA® MMR live attenuated vaccine (derived from Enders’ attenuated Edmonston strain) and RNA from clinical samples collected during an outbreak in South Africa, as positive controls for MeV pan and vaccine-specific primer-probe combinations respectively.

### Fever-rash surveillance and diagnostic testing

#### Fever-rash surveillance

The NICD in Johannesburg, South Africa is one of 86 accredited World Health Organization GMRLN laboratories and conducts measles testing (IgM and IgG serology, PCR, and sequencing) for South Africa and the southern Africa region[21]. Samples from suspected measles cases (any person with fever, rash and one of cough, conjunctivitis or coryza) are collected by provincial and district health department staff and transported on ice to the NICD. Serology testing is conducted using the Euroimmun Anti-Measles Virus ELISA (IgM and IgG) (Lubeck, Germany). PCR testing is conducted using the method described in Chapter 6 of the WHO Manual for the Laboratory-based Surveillance of Measles, Rubella, and Congenital Rubella Syndrome[21,22] targeting a region of the N gene. South African surveillance indicators have consistently achieved more than the required two non-measles fever-rash cases/100,000[23].

When vaccination history is not provided (most specimens submitted for testing), NICD surveillance protocols define a laboratory-confirmed case of measles as a positive IgM serologic test for MeV in a person with fever, rash and one of cough, conjunctivitis or coryza. We extracted measles serology results (including district where sample was collected and collection date, without identifying data) for the period 16 February 2021 (week 7) to 08 March 2024 (week 10) from our NICD Centre for Vaccines and Immunology database by district of South Africa on 29 March 2024.

### Data analysis

#### Comparison of dPCR and qPCR results, determination of limit of detection and normalisation of data

We conducted univariable regression analysis using MS Excel comparing qPCR Ct thresholds and dPCR genome copies/reaction for each dilution of the positive control in monoplex (MeV) and duplex (MeV plus RuV) assays to determine the limit of detection (LoD) and to assist with interpretation of low concentrations of MeV detected by dPCR. We normalised our data by determining the ratio of MeV:PMMoV genome copies per microlitre.

#### Comparison of clinical surveillance and wastewater measles testing data at health district level by week

As public health responses to detection of measles cases are triggered by the number of cases detected in a health district over one month[24], we elected to compare wastewater and clinical surveillance data by health district. Further, we temporally limited each comparison to a single epidemiological week, and, in a second analysis, to the epidemiological weeks before and after a week when a clinical case was detected to accommodate for the period during which wild-type MeV is excreted[25]. We conducted this analysis as follows: we grouped and tallied the total number of IgM-positive clinical specimens submitted for testing by epidemiological week of sample collection and district of health facility where the case was identified using case-line lists collected as part of national measles fever-rash surveillance. We also grouped and tallied wastewater samples by epidemiological week and district. We merged these clinical and wastewater results by epidemiological week and district and eliminated week-district pairs where no wastewater samples were tested.

We defined a ‘positive concordant wastewater-clinical pair’ as any instance in a given epidemiological-week in a specified district where at least one wastewater sample tested positive for MeV and one clinical case was identified. Conversely, we defined a ‘negative concordant pair’ as one where all wastewater and clinical samples tested negative or no clinical samples were submitted. The remaining ‘discordant’ pairs were those where least one case was detected but all wastewater samples were negative, or vice versa. We determined and described the proportion of concordant and discordant week-district pairs and presented these in two-by-two tables. In concordant instances, we also determined the relationship between the number of measles cases detected and the proportion testing positive as well as the number of measles cases detected and the wastewater levels of MeV in genome copies/mL using simple regression (MS Excel). When MeV was detected in wastewater, but no clinical cases were detected in that district, we examined the clinical measles surveillance data in neighboring districts. To do this, we summed, tabulated and visually inspected the measles cases from neighboring districts in the same epidemiological week, and the weeks before and after.

### Ethics

The protocol for this study was reviewed and approved by the University of the Witwatersrand Human Research Ethics Committee (HREC) (MM220904). The National Department of Health’s Environmental Health and Expanded Programme of Immunisation directorates, which are responsible for coordinating polio environmental surveillance, arranged permission from the municipal departments of water and sanitation for the NICD to test wastewater from participating wastewater treatment plants sites for polio, SARS-CoV-2 and other vaccine-preventable diseases. The City of Tshwane, City of Ekurhuleni and City of Johannesburg gave permission to collect and test wastewater samples from inspection holes located in their regions. Measles surveillance data is collected by the NICD as part of the institution’s responsibility to conduct national public health surveillance, in accordance with the oversight provided by the University of the Witwatersrand’s HREC (M210752)

## Results

### RT-dPCR assay optimisation and specificity

We optimized the QIAcuity OneStep Advanced Probe RT-dPCR assay by increasing the manufacturer’s recommended cycling profile of 30s and cycle number of 40 to one minute and 45 cycles respectively to obtain optimal separation of positive and negative partitions on the RT-dPCR platform.

MeV pan primers detected every instance of spiked GMRLN positive control, spiked MMR live-attenuated vaccine and spiked MeV in known-MeV positive clinical samples collected during an outbreak in South Africa (Table 2). However, in-house genotype A-specific primers falsely detected vaccine strain when only wild-type strain was present in 2 of five instances (Table 2). Primer-probes designed to detect wild-type genotypes recently circulating in South Africa (B3, D8 and H1) were non-specific as they intermittently detected genotype A and lacked sensitivity (Table 2).

**Table 2.** Results of MeV (pan), genotype A and wild-type (genotype B3, D8 and H1)-specific primer-probe combinations when tested using appropriate controls*.

| Primer probe combination | Type of positive control (#positive /# tested) |  |  |
| --- | --- | --- | --- |
|  | Vaccine | CDC MeV positive control | Wild-type |
| MeV pan | 5/5 | 5/5 | 5/5 |
| MEV - genotype A | 5/5 | 5/5 | 2/5<sup>#</sup> |
| MeV - WT | 5/5<sup>*</sup> | 5/5 | 2/5<sup>1</sup> |

\*Raw data in Table S1 of the supplement
\*These results indicate that the WT PCR reactions are positive when only measles vaccine (genotype A) is present. Therefore MeV-WT PCR reactions are non-specific
<sup>#</sup>These results indicate the genotype A primers are positive in some instances when only WT is present. Therefore, the genotype A PCR reactions are non-specific
<sup>1</sup>These results indicate that wild-type primers were positive in only some instances when wild-type is present. Therefore MeV-WT lack sensitivity

### Assay limit of detection

Following optimisation of qPCR and dPCR protocols for MeV pan and MeV pan multiplexed with RuV, we determined the relationship between the number of positive dPCR partitions and Ct values of qPCR (Figure 3). The limit of detection in duplex reaction with RuV was determined to be 0.346 -0.494 gc/uL (1 positive partition in assays with 100% (8500/8500) valid partitions, or 1 positive partition in assays with 70% valid partitions (5950/8500) (Figure S1).

**Figure 3.**
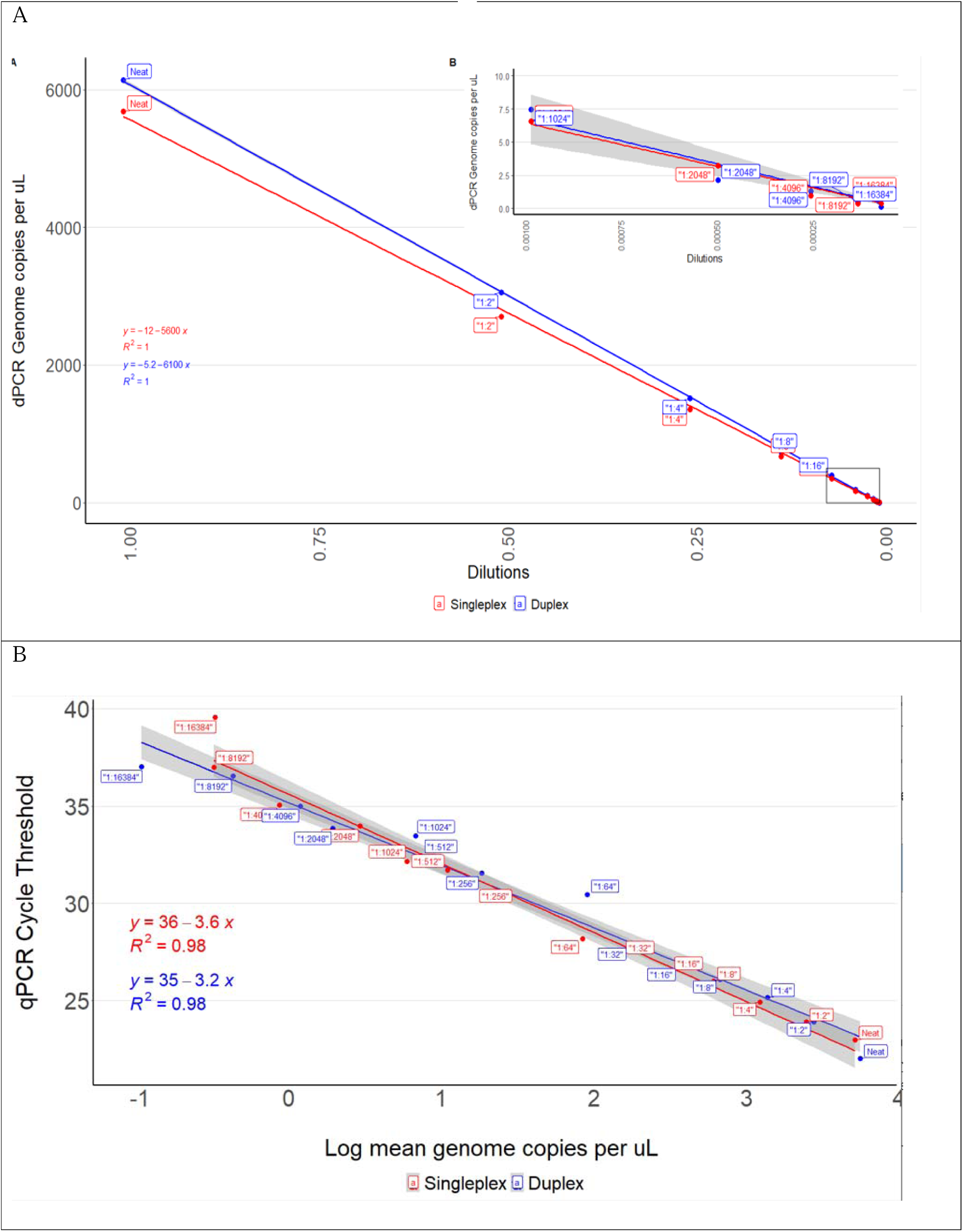
Validation of dPCR and qPCR assays (red = measles virus (MeV); blue=MeV duplexed with rubella virus (RuV)) using primer-probe combinations tested on serial dilutions (1:32 to 1:16,384) of WHO Global Measles and Rubella Laboratory Network MeV positive control sequences. Figure 3A. dPCR results in genome copies per microlitre of reaction by dilution of MeV positive control, (dilutions 1:32, 1:64, 1:128, 1:256, 1:512 omitted from the insert). Figure 3B. Cycle threshold (vertical axis) by log of genome copies per microlitre (gc/uL) as determined by qPCR.

### Testing of retained concentrates from wastewater samples

A total of 2149 wastewater concentrates had been stored after processing of grab samples collected across nine provinces between 16 February 2021 (week 7) and 08 March 2024 (week 10). Limpopo, Mpumalanga, North West and the Northern Cape provinces accounted for only 58 samples (2.6%) as sampling from these sites commenced late or ceased early (see Table 3). Retained concentrates underwent extraction and duplex dPCR in batches between November 2023 and March 2024 (RuV results not presented here). Of the total, 43 samples (2%) tested positive for MeV (Table 3). The majority of wastewater samples were collected in Gauteng province (n=1492), which also had the highest percentage of samples positive for MeV (31, 2.1%, Table 3). No MeV was detected in wastewater from the provinces with the lowest numbers of tested samples (Table 3). Over the three-year period, the majority of positive samples were identified in 2024 (Figure 4, Figure S1). In total, 103 wastewater samples were collected during the vaccination campaign, of which 4 (3%) tested positive for MeV. The median MeV concentration amongst positive samples was 2.12 (2.04-2.25) gc/mL. Pepper mild mottle virus was detected in all wastewater samples at a median concentration of 441 (276-1528) gc/mL. The median ratio of MeV:PMMoV was 441×10^-3^ (range 276×10^-3^ -1528×10^-3^).

**Figure 4:**
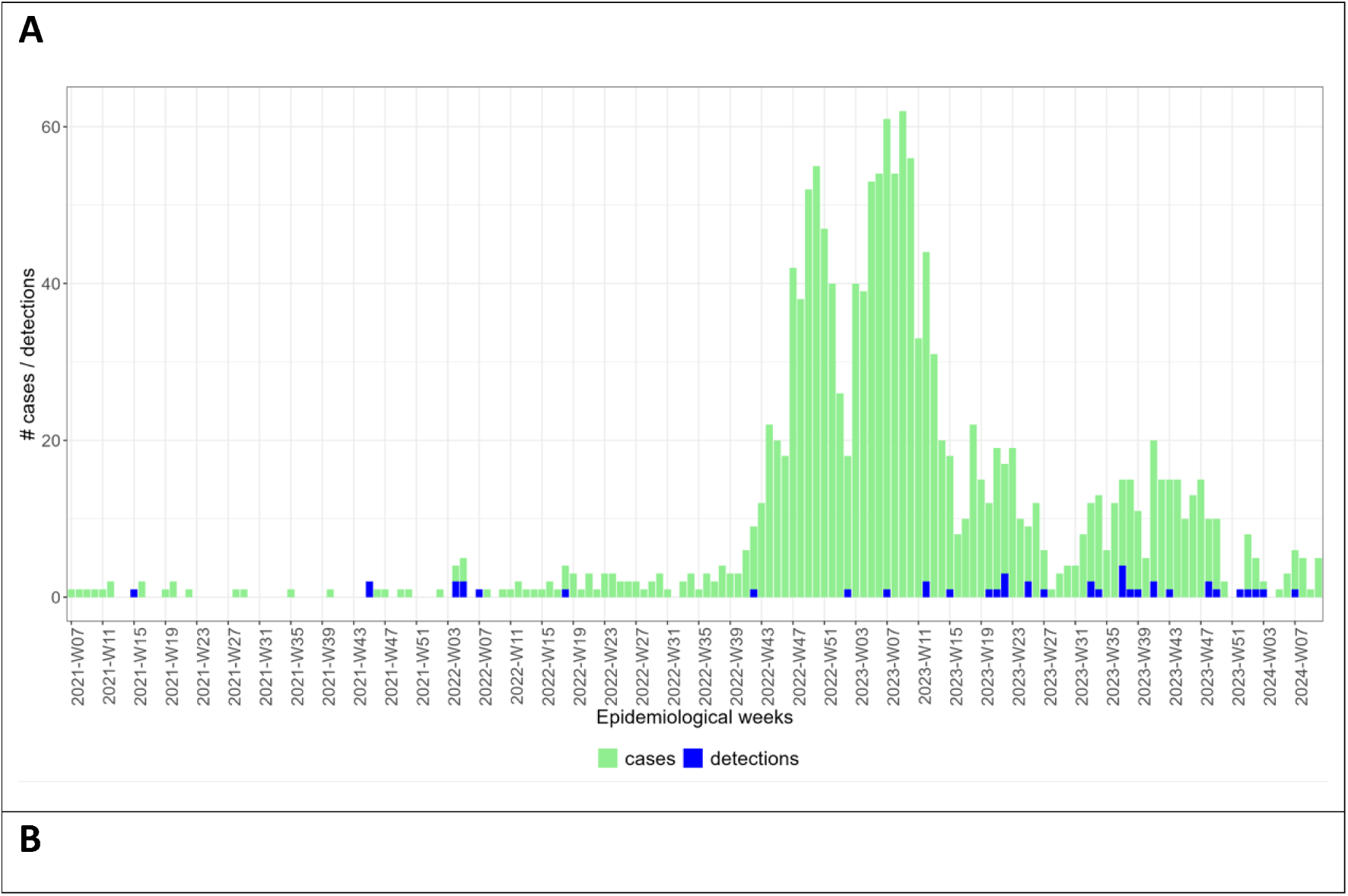
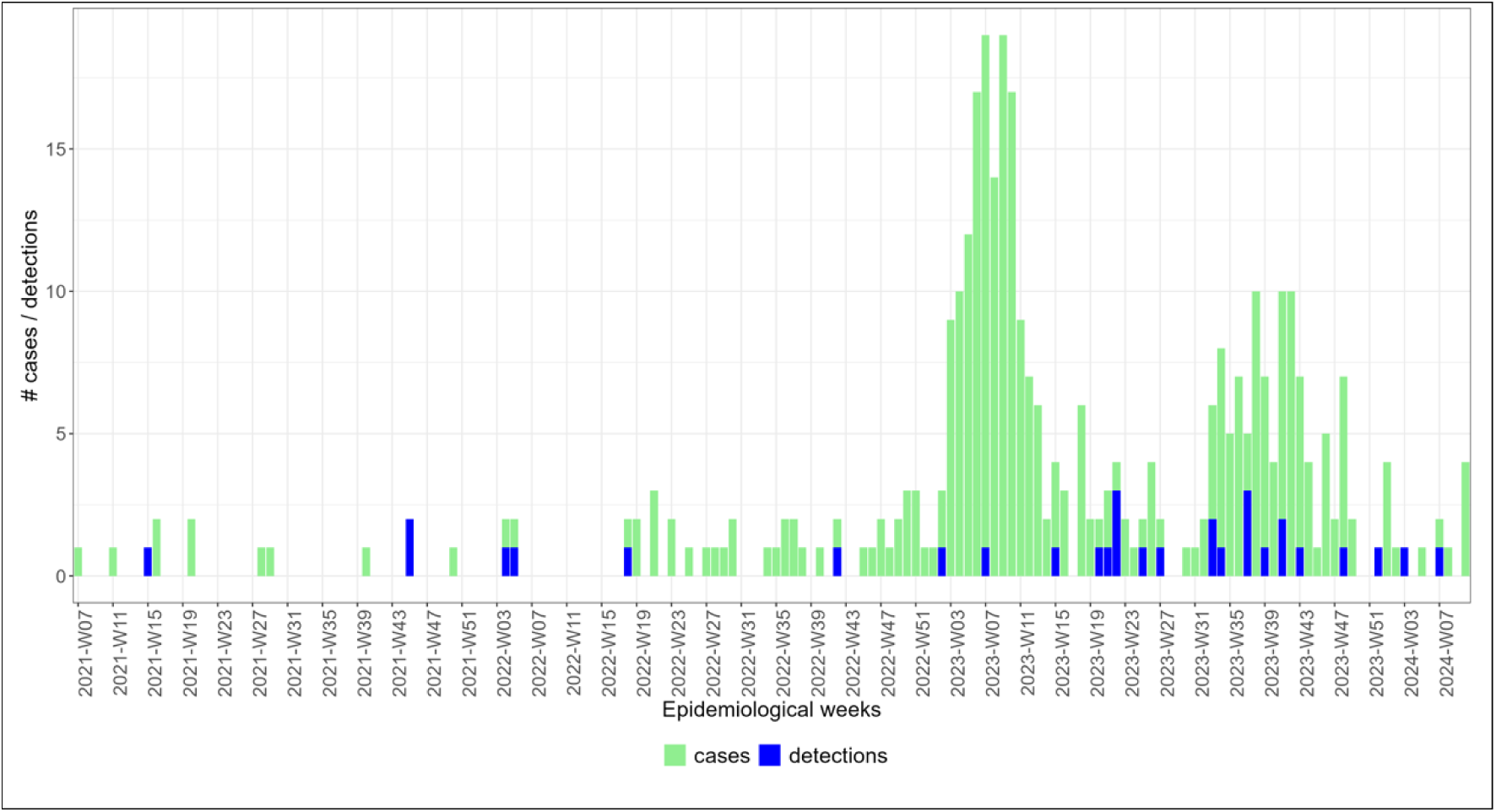
Epidemiological curve showing the number of laboratory- confirmed measles cases (measles IgM positive) submitted to the National Institute for Communicable Diseases as part of fever-rash surveillance (green bars) and the number of wastewater samples in which measles virus was detected by digital PCR (blue bars) by epidemiological week (from 2021 week 7 to 2024 week 10) from A) South Africa, and B) Gauteng Province.

**Table 3.** Wastewater samples tested for measles virus by digital PCR April 2021 – March 2024 by province and by district of Gauteng Province. The median concentration (and inter-quartile range) of measles virus (MeV) is reported in genome copies/uL from all positive samples. Quantitative results for pepper mild mottle virus (PMMoV) are reported similarly though all samples tested positive for PMMoV.

| Province | District | Time period | | Samples tested (N) | MeV detected (n, %) | MeV concentration in gc/mL (median, IQR) | PMMoV concentration in gc/mL (median, IQR) | Ratio MeV: PMMoV $\times 10^{-3}$ (median, IQR) |
| --- | --- | --- | --- | --- | --- | --- | --- | --- |
|  |  | Start | End |  |  |  |  |  |
| Eastern Cape | Buffalo City and Nelson Mandela Metro | April 2021 | March 2024 | 187 | 3 (1.6) | 2.12 (2.05-6.4) | 404 (346-504) | 7.37 (5.33-16.9) |
| Free State | Mangaung | August 2021 | March 2024 | 183 | 3 (1.6) | 2.17 (2.07-4.12) | 310 (155-548) | 11.17 (6.96-15.37) |
| Gauteng | City of Tshwane | April 2021 | March 2024 | 637 | 13 (2) | 2.12 (2.08-6.11) | 1017 (338-2871) | 3.27 (1.11-8.04) |
| Gauteng | City of Ekurhuleni | February 2021 | March 2024 | 547 | 13 (2.4) | 2.11 (2.04-2.19) | 442 (316-1421) | 5.54 (1.43-15.78) |
| Gauteng | City of Johannesburg | April 2021 | March 2024 | 308 | 5 (1.6) | 2.14<br>(2.11-2.16) | 177<br>(123-199) | 11.29<br>(10.96-17.9) |
| KwaZulu-Natal | eThekweni | August 2021 | March 2024 | 149 | 5 (3.4) | 2.04<br>(2.04-4.17) | 1079<br>(387-2343) | 4.08<br>(0.87-5.27) |
| North West | Bojanala | November 2023 | March 2024 | 22 | 0 | - | 1231<br>(653-1914) | - |
| Northern Cape | Namakwa | August 2021 | May 2022 | 14 | 0 | - | 927<br>(505 - 105) | - |
| Western Cape | City of Cape Town | August 2021 | March 2024 | 72 | 1 (1.4) | 2.1 | 1934 | 1.1 |
| Mpumalanga | Ehlanzeni | February 2024 | March 2024 | 10 | 0 | - | 616<br>(217- 821) | - |
| Limpopo | Mopani | February 2024 | March 2024 | 12 | 0 | - | 370<br>(201 - 740) | - |
| South Africa |  | February 2021 | March 2024 | 2149 | 43 (2) | 2.12<br>(2.04-2.25) | 441<br>(276-1528) | 5.27<br>(1.7-15.72) |

MeV=measles virus, PMMoV-pepper mild mottle virus, IQR=inter-quartile range

### Comparison of clinical surveillance and wastewater measles testing

We identified 481 epidemiological week-districts where both wastewater samples and clinical diagnostic tests were submitted for surveillance. Amongst week-districts, some districts had multiple wastewater samples (median=2, range 1-19 samples) for a given week from a number of sampling locations (n=1-7), while 357 week-district pairs had more than one positive clinical case per week, ranging from 1-104 cases, with a median of 3 cases per week.

Amongst all week-district pairs, measles was identified by both wastewater and clinical surveillance in 2.9% (14) pairs and by neither wastewater nor clinical surveillance in 343 (71%) of pairs (Table 4). Discordance was observed in the remaining 124 (25.8%) of week-district pairs of which the larger proportion (23.1%) were those with negative wastewater samples in districts where clinical cases were detected. When the time frame for a concordant positive test was broadened to include the presence of clinical cases up to a week before or after a positive wastewater sample, the number of positive concordant pairs increased to 17 (3.5%), whilst the number of time-district pairs where wastewater detected evidence of measles and clinical surveillance failed to detect a case, decreased to 10 (2.1%, Table 4). No dose-relationship was discernable between the concentration of MeV detected in wastewater and the number of laboratory-confirmed cases in the same district and epidemiological week (Figure S2). Amongst discordant week-district pairs where MeV was detected in wastewater (Table 5), no clinical testing was conducted in nine districts during that week, whilst 1-15 laboratory-confirmed cases of measles were identified in eight of 14 neighbouring districts.

**Table 4.** A comparison of wastewater and clinical testing results in the same district and epidemiological week for surveillance conducted between epidemiological week 7, 2021 to epidemiological Week 10, 2024.

| **Part A: same epidemiological week** |  | **Wastewater Results** |  |  |
| --- | --- | --- | --- | --- |
|  |  | Positive | Negative | Total |
| Clinical Results (IgM) | Positive (IgM) | 14 (2.9%) | 111 (23.1%) | 125 |
|  | Negative (IgM) | 13 (2.7%) | 343 (71.3%) | 356 |
|  | Total | 27 | 454 | 481 |
| **Part B: same epidemiological week + clinical results one week before and one after** |  | **Wastewater Results** |  |  |
|  |  | Positive | Negative | Total |
| Clinical Results (IgM) | Positive (IgM) | 17 (3.5%) | 190 (39.5%) | 207 |
|  | Negative (IgM) | 10 (2.1%) | 264 (54.9%) | 274 |
|  | Total | 27 | 454 | 481 |

**Table 5.**
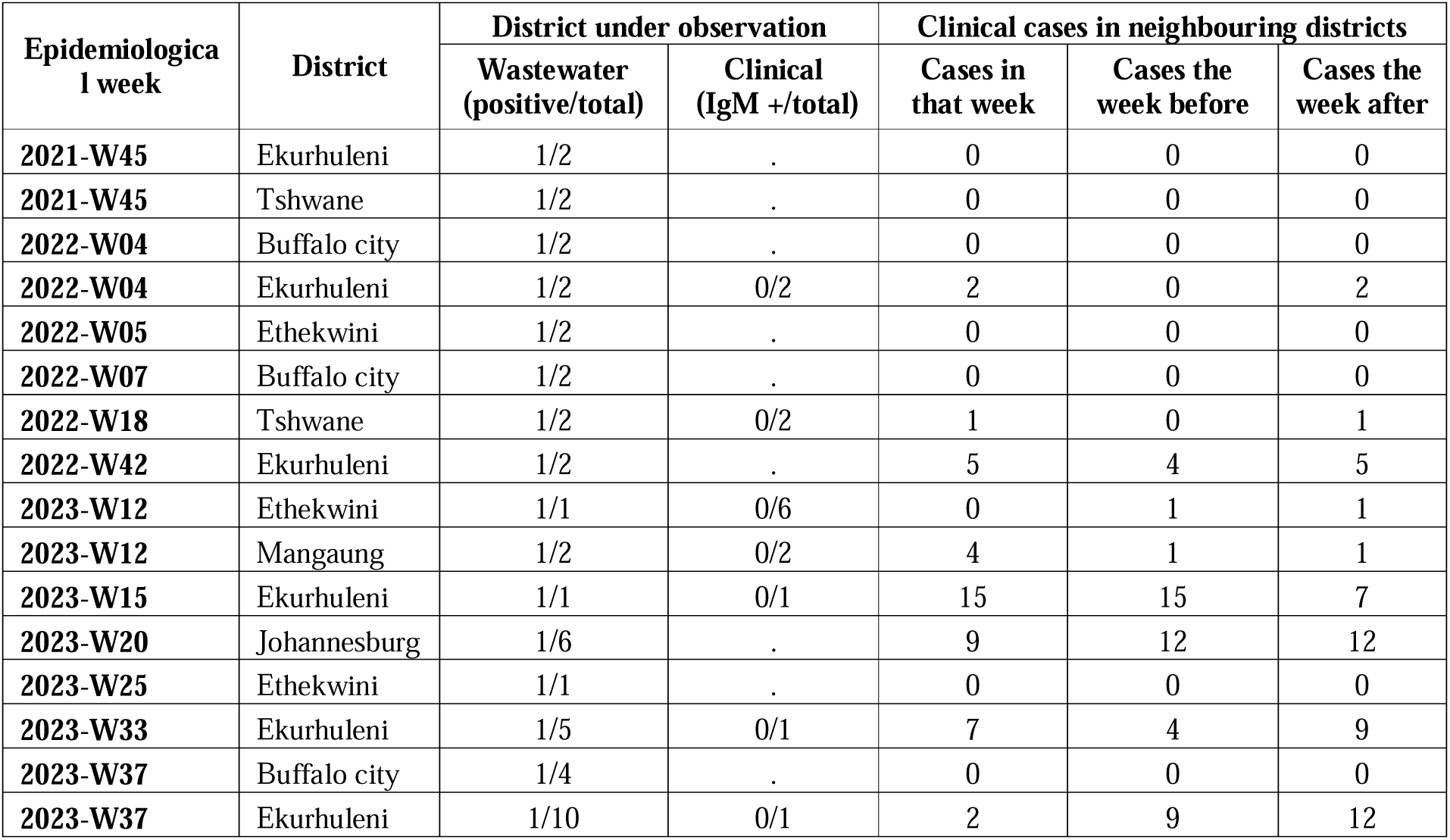
Fever-rash surveillance results for epidemiological weeks and districts where wastewater samples tested positive for measles RNA during epidemiological week 7, 2021 to epidemiological Week 10, 2024. Epidemiological weeks are designated with the year followed by the week number.

## 4. Discussion

We successfully developed and validated a duplex dPCR for the detection of measles virus in wastewater. On application to retained wastewater concentrates sampled from national and sentinel site surveillance during an ongoing measles outbreak in South Africa, the assay detected MeV RNA at copy numbers ranging from 1.97 – 165.8 gc/mL in 43 (2%) of wastewater samples. Despite likely RNA decay on account of sample storage, in an analysis based on district-time pairs, we observed positive MeV wastewater detections in health districts where no clinical cases were detected. This observation, together with recent global results of MeV WES testing during outbreaks illustrate that wastewater surveillance should be explored using real-time sampling and strengthened laboratory test methods for MeV, to explore how data emerging from MeV WES may complement clinical fever-rash surveillance for measles cases, and if MeV WES may have value as a programmatic surveillance tool.

We believe these detections of MeV in wastewater are representative of measles infections in the catchment area and health district, and that this has public health significance, despite the low concentrations of MeV in our samples. Firstly, it is to be expected that MeV will be present in wastewater as the virus is excreted in urine by infected persons [25,26]. Further, grab sampling, despite being an instantaneous sample, may provide useful results even if few clinical cases are present in a sewershed because dispersal of virus occurs the sewerage network due to differing gradients, varying flow rates and entrapment in sediment[27]. Our findings are not unique – even though we found MeV in low copy numbers, we know this is likely because of the well-established loss of RNA following freeze-thaw cycles [14,28], and even with real time sampling, other authors have also detected MeV in wastewater in an outbreak context in low copy numbers ranging from 2-22gc/mL[9,13,15,16]. Our analytic approach further demonstrates the public health significance of our findings, in that by comparing wastewater and clinical surveillance data at a district level, and by epidemiological week, we provide results to support the WHO definition of a measles outbreak (a threshold of 2-5 cases *per district* in one month is required before initiating appropriate public health interventions[24,29]), and identify instances where measles transmission is evident despite the absence of clinical cases. Programmatic fever-rash and laboratory-based surveillance for measles has never had a way to demonstrate surveillance sensitivity, because there was no way of demonstrating MeV transmission where case detection is limited by patient health seeking behaviour, health system issues and clinician failure to submit laboratory tests for measles. Detractors may argue that wastewater surveillance also lacks sensitivity, as in our study, wastewater surveillance was not always positive in districts where laboratory-confirmed measles cases were detected. This is likely due to the intrinsic limitations of wastewater detection for viral RNA targets, such as viral decay in wastewater matrix, and/or excessive dilution, or timing of sampling, but also geographical mismatch between case location and wastewater catchment area, and RNA decay during storage and freeze-thaw cycles. However, these limitations do not detract from the value of positive wastewater detections that highlight deficiencies in clinical surveillance programmes. Put together, the public health value of measles wastewater surveillance lies in its ability to detect active measles transmission where clinical surveillance fails to detect cases.

In the light of the public health significance of measles WES, consideration may be given to potential roles of WES in measles control and elimination. In areas where fever-rash surveillance is weak or absent, as may be the case in resource poor settings, wastewater surveillance may indicate the presence and extent of community transmission of MeV. Where measles is not endemic, or measles outbreaks are not ongoing and especially where vaccination coverage is less than 95%, wastewater detections of MeV may provide early detection signals that herald the onset of outbreaks. The potential for sequencing of MeV from wastewater has been demonstrated [30], and the genomic analyses that are conducted may be useful to complement data obtained from sequencing clinical strains, or to provide genomic data in resource-poor contexts where samples appropriate for molecular detection and sequencing (throat swab or urine) are not submitted for testing. The additional genomics data and phylogenetic analysis obtained from wastewater-derived sequences will strengthen measles control in the same way that environmental surveillance has strengthened polio control, particularly if whole genome MeV sequencing replaces the n450 genotyping methods currently in use[31].

Lastly, if WES for measles is to be included in programmatic control, public health interventions that should follow detection of MeV in wastewater need to be established. Following detection of certain polio strains in wastewater, the WHO guidelines require district-wide SIAs. Following detection of laboratory-confirmed clinical measles cases, WHO presently recommends case investigation and diagnostic testing, contact tracing with ring vaccination, case finding, and district wide supplementary immunisation activities (SIA) if a threshold of cases (two to five cases per health district per month) are identified[24]. WES measles detection will not facilitate case identification but may identify vulnerable communities where clinical surveillance should be strengthened. As the sensitivity of WES MeV surveillance becomes clearer, it may become evident if WES MeV detection should trigger a supplementary immunization activity.

Lastly, our findings were limited by likely RNA decay discussed above, and by our inability to design primers and probes with sufficient sensitivity and specificity to differentiate MeV-WT from vaccine strain (MeV genotype A). This meant that we were unable to rule out the presence of genotype A, and to rule out the presence MeV-WT if genotype A was detected. This is not surprising given the marked similarity between genotypes A, B3, D8 and H1 in the N gene region selected for primer design. Even when applied to clinical samples, differentiating WT from vaccine without sequencing is challenging, often relying on probes that differ by a single nucleotide[32,33]. Under these circumstances, differences in Ct thresholds between MeV-WT and vaccine-specific probes rather than presence/absence signals are helpful to distinguish the two[34]. In a similar way, Wu et al[14] successfully achieved WT vs vaccine differentiation in wastewater samples using primer-probe combinations targeting the M gene. Their format relies on differences in dPCR amplitudes (indicative of degree of fluorescence) emanating from positive partitions to differentiate WT from vaccine, relying on the principle that mismatched probes release less fluorescent dye following incomplete breakdown by the exonuclease function of Taq polymerase. Our laboratory is presently testing the Wu method in parallel with routine testing of wastewater for MeV.

However, vaccinated, non-immune persons secrete lower levels of vaccine virus and for a shorter duration, leading to lower inoculum sizes in wastewater. Therefore, it is much more likely that MeV detected in wastewater is wild type strain, so even without the ability to differentiate WT-MeV from vaccine strain, wastewater detection remains useful. The evidence base for this hypothesis is as follows: Firstly, no onward transmission of vaccine strain has ever been documented suggesting that vaccine excretion rates are low[35]. Shedding duration in WT measles cases may be as long as 26-61 days post infection[26], whilst Walsham et al reported median PCR detection of measles vaccine in urine post MMR vaccination of 11 days, (IQR 9,2 days)[36]. Lastly, routine EPI measles vaccination is administered to infants who are in diapers and may not contribute to wastewater, and even during a measles SIA, most vaccine recipients have seroprotective antibodies that prevent viraemia.

A further laboratory consideration and potential limitation of our work is the lack of clarity regarding the need for a normalization step to MeV detection results. Normalisation may remove apparent fluctuations in pathogen concentration when changes in wastewater volume (e.g. through rain) occur and may allow for elimination of ‘noise’ or false signals. However, in an elimination context, as with polio, pathogen concentrations are of less significance compared to detection results. Nonetheless, we elected to use PMMoV concentrations in wastewater, a reliable marker of fecal load. The ratios of MeV:PMMoV were largely constant across sewersheds, differing only by 1 log, and did not reflect the large increase in measles cases during the 2022-23 outbreak, likely because RNA decay in our retained concentrates may have hidden this signal.

Finally, our analysis strategy and positive findings, even though only a small percentage of wastewater samples tested positive, together suggest that real-time WES for measles holds promise to support the global measles and rubella elimination agendas. Evaluation of more efficient wastewater concentration, extraction and detection methods for MeV to improve sensitivity and provide sufficient material for genotyping are desirable. Measles WES with real-time wastewater sample processing should be integrated into clinical measles surveillance programmes in routine contexts to accumulate experience and data to allow for evaluation of the relative contribution of this novel surveillance modality to measles control and elimination.

## Supporting information

Epidemiological data - week-district pairs

## Data Availability

ll relevant data are within the manuscript and its Supporting Information files.
2] Epidemiological data pertaining to the number of clinical cases per epidemiological week and district, together with the wastewater data is uploaded along with this manuscript.

## Acknowledgements

We would like to acknowledge the contribution of the NICD Centre for Vaccines and Immunology for use of the polio environmental laboratory, and measles surveillance data (also approved by Wits HREC). We thank the municipal workers, and NICD drivers who collected and transported certain wastewater samples. We thank our sample collectors Lebohang Rabotapi, Lethabo Monametsi, and laboratory technicians Thabo Mangena, Mantshali Motloung for collecting and initial processing of the samples, and our administrator Namhla Madikane for her helpful support and willingness to go to the field when necessary. Thank you to Dr Colleen Bamford for attention to detail and finalizing the manuscript.

## Supplementary material

