## Supplement for "Wastewater testing during the South African 2022-2023 measles outbreak demonstrates the potential of environmental surveillance to support measles elimination"

**Table S1: Sensitivity and specificity of RT-dPCR for detection and differentiation of measles wild-type and vaccine genotypes**

| Well | Control | Target (Name) | Conc. [cp/ÂµL] (dPCR reaction) | CI (95%) (dPCR reaction) | Partitions (Valid) | Partitions (Positive) | Partitions (Negative) |
| --- | --- | --- | --- | --- | --- | --- | --- |
| A5 | CVI06101/24 (WT) | MV_vaccine | 0 | - | 8251 | 0 | 8251 |
| A5 | CVI06101/24 (WT) | MV_wildtype | 0 | - | 8251 | 0 | 8251 |
| B5 | CVI06272/24 (WT) | MV_vaccine | 2,423 | 80,00% | 8267 | 6 | 8261 |
| B5 | CVI06272/24 (WT) | MV_wildtype | 2,827 | 74,10% | 8267 | 7 | 8260 |
| C5 | CVI06937/24 (WT) | MV_vaccine | 0 | - | 8254 | 0 | 8254 |
| C5 | CVI06937/24 (WT) | MV_wildtype | 0 | - | 8254 | 0 | 8254 |
| D5 | CVI06282/24 (WT) | MV_vaccine | 0 | - | 8262 | 0 | 8262 |
| D5 | CVI06282/24 (WT) | MV_wildtype | 0 | - | 8262 | 0 | 8262 |
| E5 | CVI06562/24 (WT) | MV_vaccine | 1,685 | 98,00% | 8242 | 4 | 8238 |
| E5 | CVI06562/24 (WT) | MV_wildtype | 2,106 | 87,70% | 8242 | 5 | 8237 |
| A6 | MeV CDC control | MV_vaccine | 30,67 | 22,20% | 8258 | 78 | 8180 |
| A6 | MeV CDC control | MV_wildtype | 31,46 | 21,90% | 8258 | 80 | 8178 |
| B6 | MeV CDC control | MV_vaccine | 28,73 | 23,60% | 8215 | 69 | 8146 |
| B6 | MeV CDC control | MV_wildtype | 29,57 | 23,30% | 8215 | 71 | 8144 |
| C6 | MeV CDC control | MV_vaccine | 24,52 | 25,50% | 8257 | 59 | 8198 |
| C6 | MeV CDC control | MV_wildtype | 24,52 | 25,50% | 8257 | 59 | 8198 |
| D6 | MeV CDC control | MV_vaccine | 24,57 | 26,00% | 8252 | 57 | 8195 |
| D6 | MeV CDC control | MV_wildtype | 25 | 25,70% | 8252 | 58 | 8194 |
| E6 | MeV CDC control | MV_vaccine | 22,89 | 26,70% | 8253 | 54 | 8199 |
| E6 | MeV CDC control | MV_wildtype | 27,99 | 24,10% | 8253 | 66 | 8187 |
| A7 | Vaccine | MV_vaccine | 257,8 | 7,80% | 8286 | 632 | 7654 |
| A7 | Vaccine | MV_wildtype | 257,4 | 7,80% | 8286 | 631 | 7655 |
| B7 | Vaccine | MV_vaccine | 272 | 7,80% | 8173 | 633 | 7540 |
| B7 | Vaccine | MV_wildtype | 272,5 | 7,80% | 8173 | 634 | 7539 |
| C7 | Vaccine | MV_vaccine | 291,2 | 7,60% | 8264 | 674 | 7590 |
| C7 | Vaccine | MV_wildtype | 289,9 | 7,60% | 8264 | 671 | 7593 |
| D7 | Vaccine | MV_vaccine | 282 | 7,70% | 8266 | 646 | 7620 |
| D7 | Vaccine | MV_wildtype | 279,7 | 7,70% | 8266 | 641 | 7625 |
| E7 | Vaccine | MV_vaccine | 299 | 7,60% | 8268 | 672 | 7596 |
| E7 | Vaccine | MV_wildtype | 297,6 | 7,60% | 8268 | 669 | 7599 |

A Eastern Cape Province

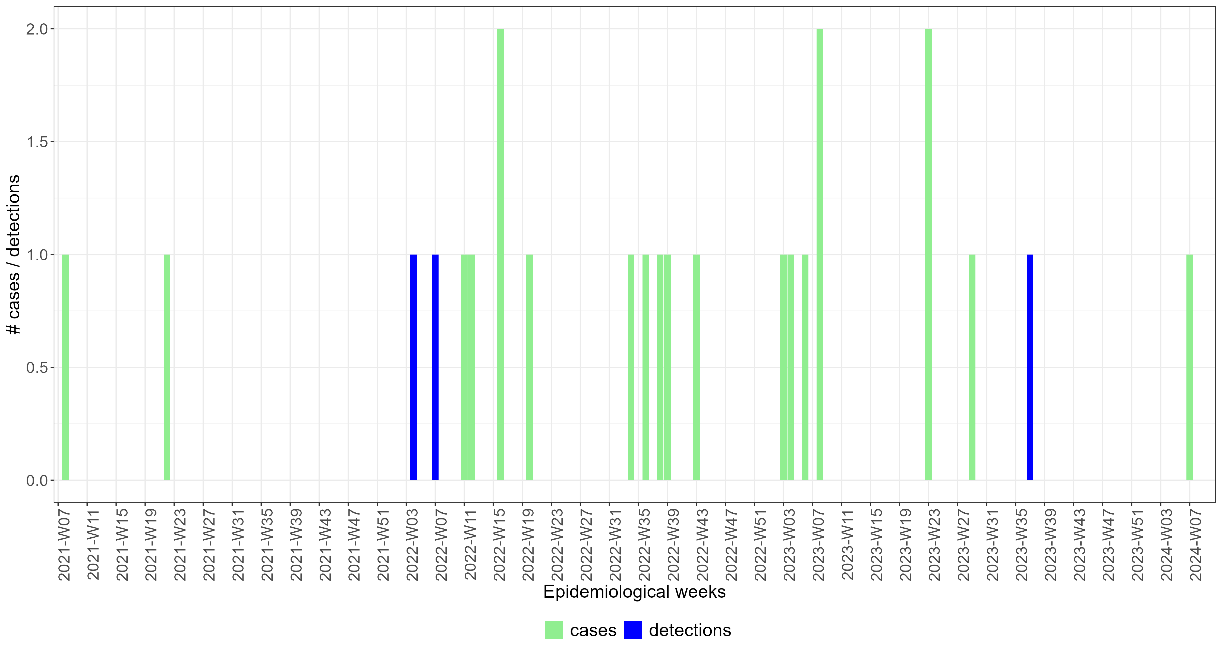

B Free State Province

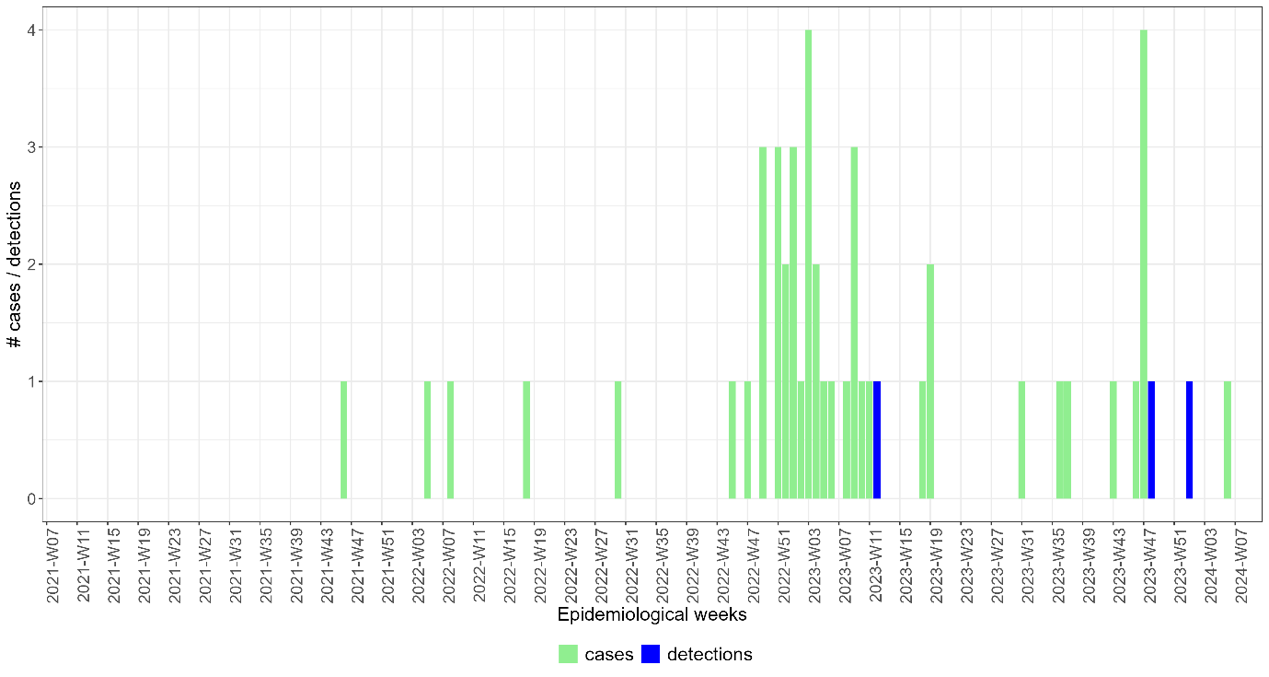

C KwaZulu-Natal Province

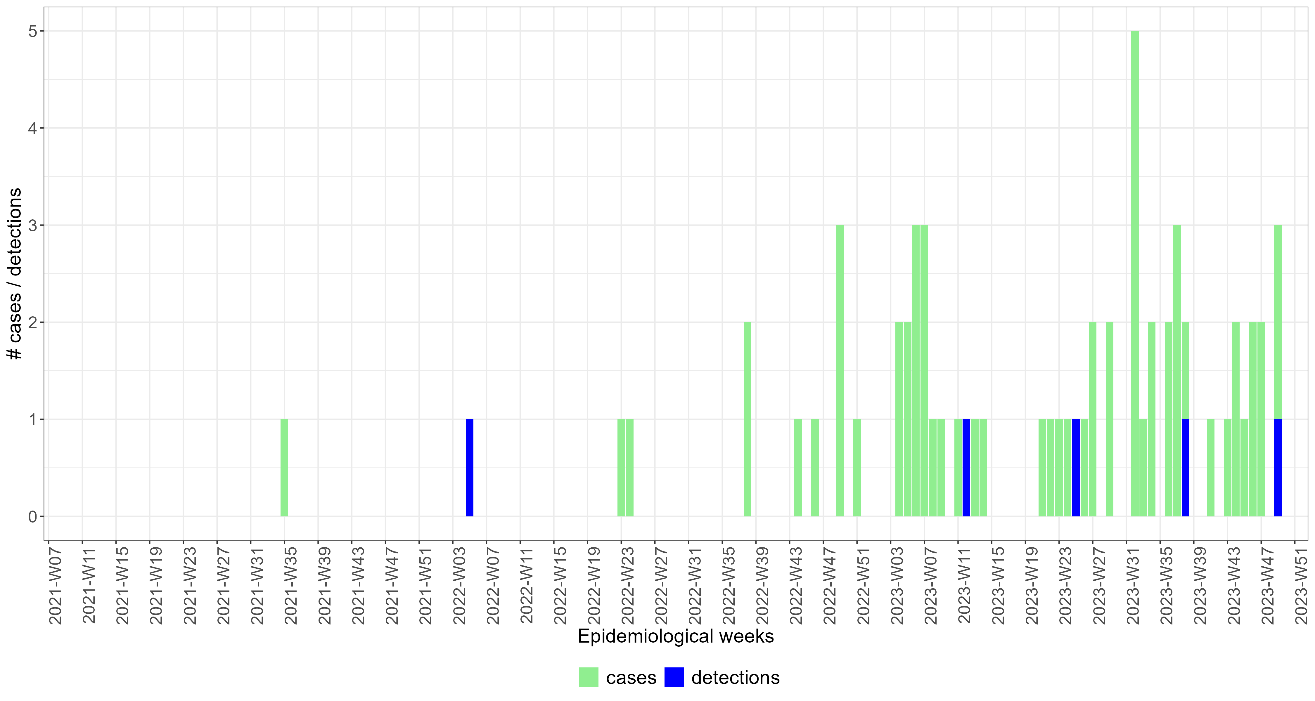

D Western Cape Province

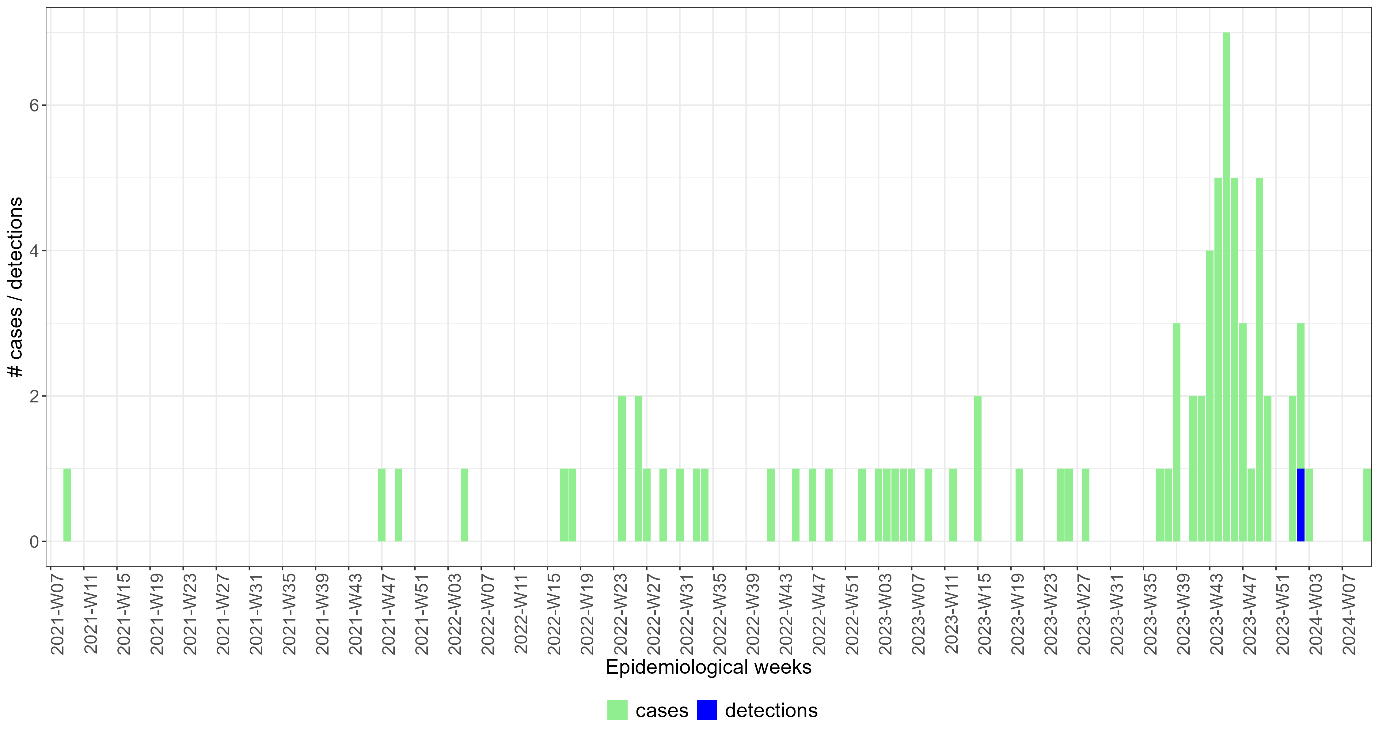

**Figure S1: Epidemiological curve showing the number of laboratory- confirmed measles cases (measles IgM positive) submitted to the National Institute for Communicable Diseases as part of fever-rash surveillance (blue bars) and the number of wastewater samples in which measles virus was detected by digital PCR (green bars) by epidemiological week (from 2021 week 7 to 2024 week 10) from four provinces in South Africa: A) Eastern Cape B) Free State C)KwaZulu-Natal D) Western Cape.**

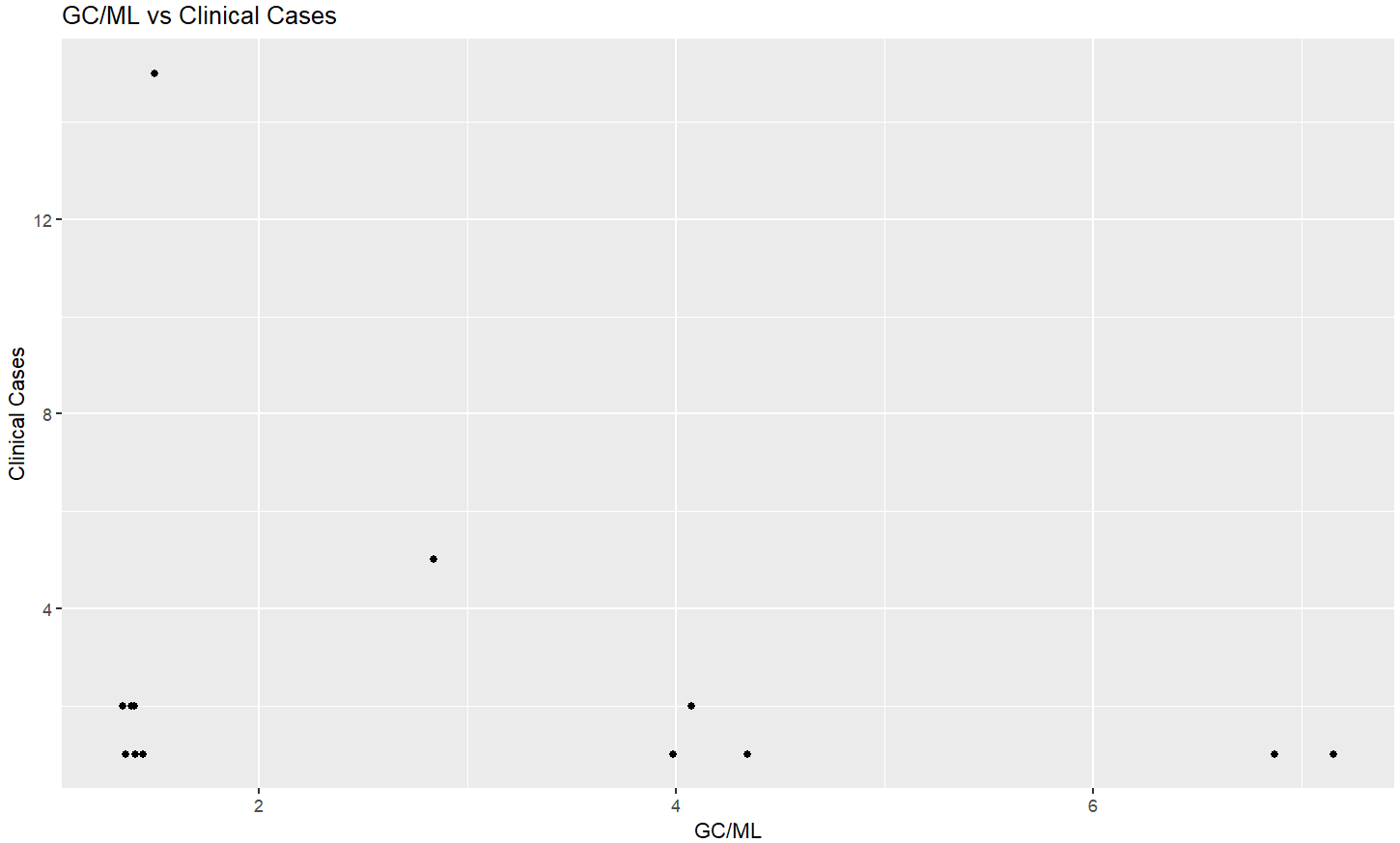

**Figure S2: The number of laboratory-confirmed measles cases (‘Clinical Cases’, vertical axis) by the number of genome copies of measles virus per millilitre of wastewater (‘GC/ML’, horizontal axis) in wastewater collected from a sample collection point in the district where the measles case was identified (n=43)**
